# Factors Associated with Acceptance of Indoor Residual Spraying (IRS) among Residents of Luwingu District, Northern Province of Zambia

**DOI:** 10.1101/2024.08.29.24312773

**Authors:** Moses Mkosha, Brown Ngenda, Mukumbuta Nawa

## Abstract

**Introduction:** Indoor Residual Spraying (IRS) is a crucial intervention for malaria control, yet its acceptability in urban areas remains understudied, particularly in Luwingu District, Zambia. This research aimed to estimate acceptance of IRS and identify the factors associated with its acceptability in an urban setting of this area that has holoendemic transmission of malaria.

**Methods:** A cross-sectional study was conducted using a structured questionnaire administered to 344 households. Data was analysed using the Statistical Package for the Social Sciences (SPSS. 28.0) included descriptive statistics and logistic regression to identify factors associated with IRS acceptability.

**Results:** A total of 344 household heads were interviewed in this study, out of which only a third (93/344) were female. Among the surveyed households, 53.3% reported being sprayed with IRS, while 46.8% were not and the difference was not statistically significant. The analysis revealed several key factors influencing IRS acceptability. Age was found to be significant, with younger individuals more likely to accept IRS. Similarly, those in formal employment and those with good attitudes towards IRS were likely to accept IRS implementation. On the other hand, sex and marital status were not associated with IRS acceptability.

**Conclusion:** This study found that Indoor Residual Spraying in an urban community of an area that has holoendemic transmission of malaria covered only about half of the households. The factors associated with acceptance of IRS included younger age group below 35 years compared to those aged 36 years and older, those in formal employment compared to those in informal employment and those with moderate and good attitudes towards IRS. On the other hand, sex, marital status and educational levels if the heads of the households were not statistically associated with acceptance of IRS implementation in their households.

## Introduction

Indoor Residual Spraying (IRS) is a widely used intervention for vector control in malaria-endemic areas. Several studies have emphasized the importance of IRS in reducing malaria transmission rates and improving public health outcomes (WHO 2019). Indoor Residual Spraying is a critical component of malaria control strategies globally. It involves applying long-lasting insecticides to the walls and ceilings of homes to kill mosquitoes that come into contact with these surfaces. The World Health Organization (WHO) has identified IRS as one of the most effective measures for rapidly reducing malaria transmission in endemic regions (WHO, 2019).

Awareness of Indoor Residual Spraying (IRS) and its benefits is fundamental to achieving high coverage rates and ensuring effective malaria control (Magaço, et al, 2019). Green et al. (2021) further supports this view, showing that targeted awareness efforts, which make information about IRS more accessible and understandable, lead to improved participation rates and better health outcomes. In Africa, where the malaria burden is disproportionately high, awareness is crucial for improving IRS coverage (Danny, et al. 2023). Hilton et al. (2023) demonstrate that in numerous African countries, the success of IRS programs is closely tied to community awareness and understanding of the intervention’s benefits. The study found that regions with robust awareness campaigns experienced better IRS coverage and a reduction in malaria incidence. This correlation highlights the critical role of public education in enhancing the effectiveness of IRS. Rajatileka et al. (2018) add that in urban settings across Africa, addressing awareness gaps and misconceptions is essential for improving IRS uptake. They emphasize that misconceptions about IRS safety and efficacy can hinder program success, suggesting that overcoming these barriers through targeted education can significantly enhance IRS coverage and impact.

In Zambia, awareness has a direct influence on IRS coverage and effectiveness; according to Chanda et al. (2020), inadequate community awareness has been a significant barrier to achieving the desired IRS coverage in Zambia. The study points out that many communities lack sufficient knowledge about the benefits and safety of IRS, which affects their willingness to participate. To address this issue, Chanda et al. (2020) recommend increased efforts in community education and engagement. The National Malaria Elimination Program (NMEC) of Zambia aligns with these findings by emphasizing the need for effective communication strategies to enhance public understanding and acceptance of IRS. The NMEC’s National Malaria Strategic Plan 2022-2026 highlights the importance of addressing awareness and knowledge gaps as part of its broader strategy to control and eliminate malaria. This study, set in Luwingu district of Northern Province of Zambia one of the high malaria transmission areas aimed at finding the acceptability and factors associated with Indoor Residual Spraying in these high transmission settings. The study will inform policy makers on the barriers to acceptability of IRS among residents in high transmission settings in Zambia.

## METHODOLOGY

### Study Design

The study used a cross-sectional design, which elicited factors associated with IRS acceptance and barriers to acceptance.

### Study setting

This study was conducted in Namukolo Urban Clinic catchment communities of Lupili and Chelstone areas of Luwingu District. Luwingu District is in the Northern Province of Zambia about 165 kilometres west of Kasama, the provincial capital. It shares boundaries with Mporokoso in the northwest, Chilubi in the south, Samfya in the South and Kawambwa in the northwest. The district has a surface area of 8,872 square kilometres with a population density of 12.8 inhabitants per square kilometer with estimated population of 112,679 and a growth rate of 5.2% per annum (Zamstat, 2020). Malaria incidence in 2023 in Luwingu district was at 784.7 per 1000 in all ages (Luwingu HMIS 2023). Malaria cases are high because of the existence of forest cover and swampy areas that are conducive as breeding grounds for malaria mosquito vectors in the district.

### Sample determination

The sample size was calculated using the formula below;

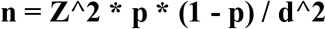

Where:

- **n** = the sample size
- **Z** = The Z score for the desired confidence level (1.96 for 95% confidence)
- **P** = estimated prevalence of the outcome (e.g., proportion of households accepting IRS) - in the absence of prior data, a value of 0.5 was used as a conservative estimate
- **d** = the margin of error, in this case 5%.

A sample size of 385 households was calculated as sufficient.

### Data Collection and Analysis

The interviews were done using a questionnaire that was administered in the local language of Bemba by trained research assistants. The data so collected was cleaned and analyzed using Statistical Package for Social Sciences (SPSS) Version 28.0. Summary frequencies, relationships and associations between variables were determined using chi-square and logistic regression. A p value of 0.05 was considered significant.

### Ethical consideration

The study adhered to ethical principles, including informed consent, confidentiality, and voluntary participation. Ethical clearance for the protocol was obtained from the Lusaka Apex Medical University Research Ethics Committee (Ethical Reference Number: 00746-24).

## RESULTS

### Basic characteristics of study respondents

A total of 344 household heads were included in this study, out of which 93 (27%) were females and 251(73%). About 53.2% (183/344) of the households were sprayed with IRS whilst 46.8% (161/344) were not sprayed; this difference was not statistically significant (P-value = 0.236). Further, there were no significant statistical differences among the respondents based on sex, age, religion, marital status, educational attainments or occupations. However, there were significant differences among the respondents based on awareness, perceptions and attitudes. Those who were aware and had good attitudes were more likely to have their households sprayed compared to those who were not aware or those with poor attitudes. Table 1 shows the summary of the basic characteristics of the respondents.

**Table 1:** Basic Characteristics of Respondents.

| Variable | Category | HH Sprayed (2022) % | HH Not Sprayed (2022) % | P-value |
| --- | --- | --- | --- | --- |
| **Sex** | Male | 123 (49.5%) | 127 (50.5%) | 0.150 |
|  | Female | 60 (63.2%) | 34 (36.8%) |  |
| **Age** | 18-25 years | 41 (56.7%) | 32 (43.3%) | 0.442 |
|  | 26-35 years | 42 (47.2%) | 46 (52.8%) |  |
|  | 36-45 years | 46 (54.3%) | 39 (45.7%) |  |
|  | Above 45 years | 54 (55%) | 44 (45%) |  |
| **Religion** | Roman Catholic | 63 (54.2%) | 53 (45.8%) | 0.245 |
|  | Seventh Day Adventist | 25 (50%) | 25 (50%) |  |
|  | Jehovah's Witness | 19 (57.1%) | 15 (42.9%) |  |
|  | United Church of Zambia | 44 (69.2%) | 19 (30.8%) |  |
|  | Pentecost | 13 (41.7%) | 17 (58.3%) |  |
|  | Others | 14 (28.1%) | 37 (61.9%) |  |
| **Occupation** | Self-employed | 37 (47.4%) | 41 (52.6%) | 0.672 |
|  | Formal employment | 96 (53.9%) | 81 (46.1%) |  |
|  | Farmers | 43 (58.9%) | 30 (41.1%) |  |
|  | Others | 10 (66.7%) | 5 (33.3%) |  |
| **Education** | Primary | 17 (58.6%) | 12 (41.4%) | 0.714 |
|  | Junior Secondary | 15 (51.7%) | 14 (48.3%) |  |
|  | Senior Secondary | 54 (51.4%) | 51 (48.6%) |  |
|  | Tertiary | 94 (52.8%) | 84 (47.2%) |  |
| **Marital** | Married | 132 (50%) | 132 (50%) | 0.321 |
|  | Single | 22 (54.8%) | 19 (45.2%) |  |
|  | Divorced | 7 (66.7%) | 3 (33.3%) |  |
|  | Widow | 22 (75.9%) | 7 (24.1%) |  |
|  | Separated | 0 | 0 |  |
| **Awareness** | Yes | 167 (66.8%) | 83 (33.2%) | <0.004 |
|  | No | 64 (56.1%) | 50 (43.9%) |  |
| **Perceptions** | Good | 34 (35.4%) | 62(64.6%) | 0.0001 |
|  | Moderate | 26 (14.9%) | 148 (85.1%) |  |
|  | Poor | 41 (55.4%) | 33 (44.6%) |  |
| **Attitudes** | Good | 60 (65.9%) | 31(34.1%) | 0.002 |
|  | Moderate | 84 (71.7%) | 33(28.3%) |  |
|  | Poor | 103 (81.7%) | 23 (18.3%) |  |

**Table 2:**
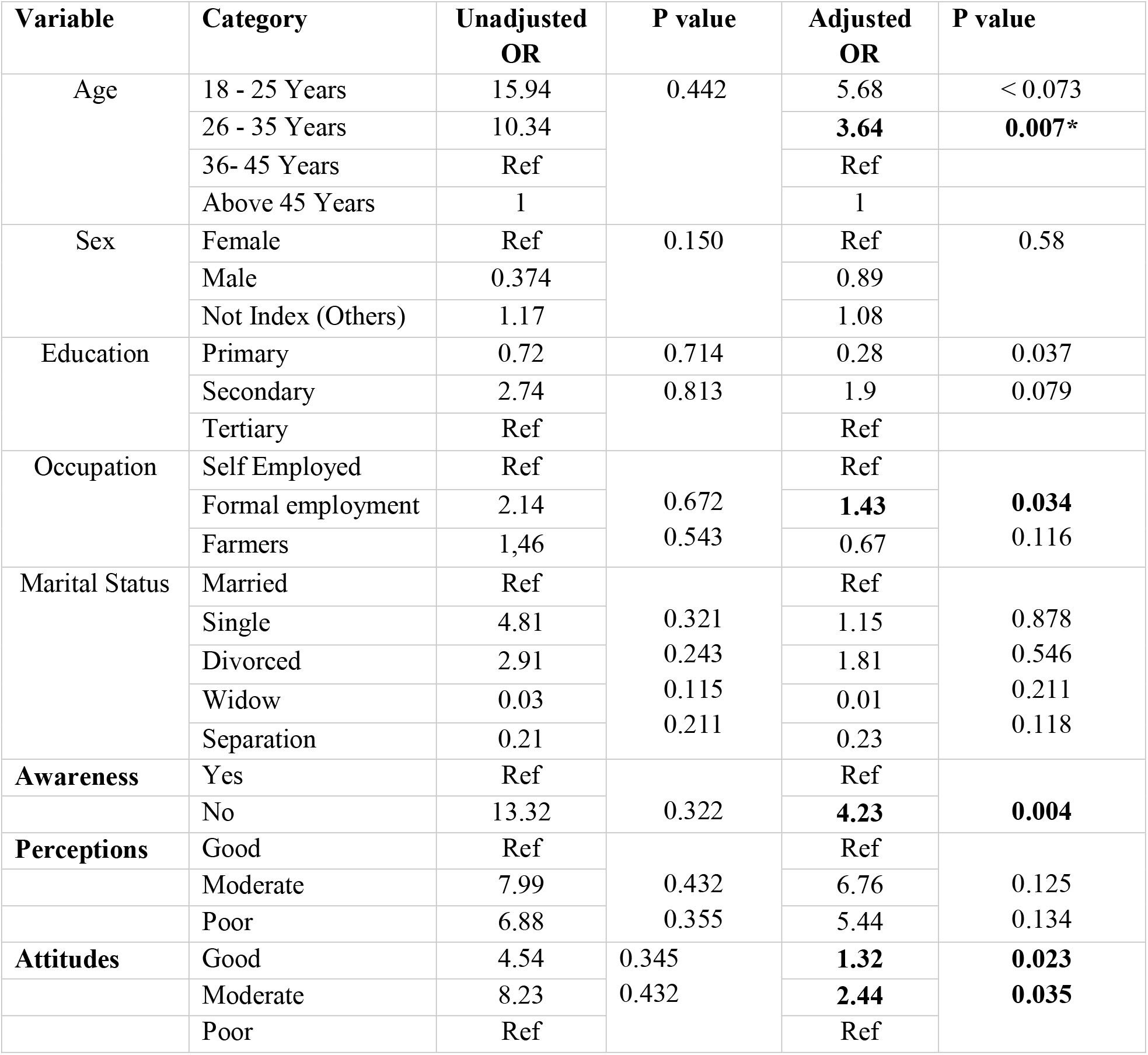
Factors associated with acceptance of IRS.

### Factors associated with Acceptance of IRS

Further analysis using logistic regression analysis examining various factors and their associations with acceptance of IRS indicated that the younger age-group of those aged 26 – 35 years were more likely to accept IRS compared to the older aged group of 36 – 45 years. In terms of education, those with less educated (primary education) aOR 0.28 (P-value = 0.037) were less likely to accept IRS compared to the reference group of those with tertiary education. Further, those with good attitudes aOR 1.32 (P value = 0.023) were more likely to accept IRS compared to those with poor attitudes. Moreover, the occupation of the head of the households were also associated with the acceptance of IRS; those in formal employment aOR 1.43 (P-value = 0.034) were more likely to accept IRS compared to those in informal employment. Ironically, those who were not aware of IRS aOR 4.23 (P-value = 0.004) were more likely to accept IRS compared to those who were aware. Other variables such as sex of the head of the household or their marital status were not statistically associated with acceptance of IRS in this study.

## DISCUSS OF FINDINGS

### Discussion

The study aimed to assess the acceptance and factors associated with the acceptance of IRS among residents in a high malaria endemic district of Luwingu in the Northern Province of Zambia. The households were equally likely to be sprayed IRS compared to those who were not sprayed and the differences in the proportions were not statistically significant. Another study in Zambia found that the overall household coverage of IRS in Northern Province in Zambia was 40.7 % down from 64.9% in 2018 while the overall national coverage of IRS in Zambia in 2021 was 39.0% (Kyomuhangi et al, 2023). This present study may have found a higher coverage of IRS in Luwingu district because it was only done in the urban residential areas of this rural district of Northern Province. The eligibility criteria for IRS is usually brick walls and iron roofs, so there more eligible structures in the research communities compared to the more rural communities of the district. Further, the drop in the coverage of IRS between 2018 and 2021 may be due to the shift in policies in 2018 and 2021, where the mass distribution of Long Lasting Insecticide-treated Nets (LLINs) in 2021 necessitated the lower need for IRS as the two interventions are no to be done in the same households. Further, the impact of COVID-19 may also have affected the deployment of interventions and resulted in lower coverage of interventions such as IRS compared to the pre-COVID-19 (Kyomuhangi, 2021). The increase in the coverage of IRS has been shown to be associated with a reduction of malaria in holoendemic malaria areas in the northern parts of Zambia (Ferris et al, 2023), the reduction in IRS coverage was therefore expected to be attended by an increase in malaria cases.

In the present study, household heads that were in the younger age group of 26 – 35 years had higher odds of refusal compared to those aged 36 and older. This is contrary to another study done in Vubwi district in Eastern Province of Zambia where age was not found to be significantly associated with refusals of IRS implementation in their households (Zhang, et al. 2024). Similarly, another study in sub-Saharan Africa in Ghana did not find age to be a factor in refusals of IRS in their households (Suuron, et al. 2020). More studies may be needed to understand the association of the age of the household head and the refusal to accept the implementation of IRS in the household in the face of conflicting findings from different areas. This study may have found a significant association with the younger age group refusing IRS possibly because younger adults in high transmission settings tend to have partial immunity to malaria and therefore their risk perception to the disease may be low and therefore may refuse to accept spraying of their households. This assertion from the authors is also corroborated by the finding that risk perception of malaria among respondents was not statistically significant (P-value = 0.125).

On the other hand, those in formal employment were less likely to accept IRS implementation in their households compared to those who were in informal employment. In a similar study done in Zambia, those who were in informal employment were less likely to accept and have their households sprayed with IRS (Zhang, et al, 2024). This finding is therefore conflicting with the present study probably because Vubwi is more rural than where the present study was done in the urban areas of Luwingu district. In more rural areas of Zambia, most housing structure are constructed with local materials such as mud walls and thatched roofs which are not suitable for IRS (Menda, et al. 2021; Nawa, et al. 2024). People in the informal sector in Vubwi who live in poor quality households may not perceive the benefit of IRS as it may not have sufficient benefits in the poor quality housing compared to modern houses which are mainly in urban settings in Zambia (Nawa, 2019).

Further, this study demonstrated that those with good and moderate attitudes towards IRS were more likely to accept it to be done in their households, however, the study also showed that being aware of IRS alone was not necessarily associated with acceptance of IRS spraying in the households. This is in tandem with the Theory of Reasoned Action and Planned Behavior, where awareness alone may not necessarily lead to the desired action but has to be attended by good attitudes towards the desired behavior to form behavioral intention and eventual action towards the desired behavior (Bui, et al, 2023). Other studies have also demonstrated that knowledge or awareness alone is not sufficient to form desired behavior in order to prevent risk factors, one study in Iran found very high knowledge of IRS but only 17% of the respondents actually had it implemented in their households (Zare, et al, 2023). Some of the barriers to acceptance of IRS include misconceptions such as IRS brings coach roaches, fleas and bedbugs, others do not want to move household properties to pave way for spray operators especially if done during the rainy season (Roberts, et al, 2021). This study therefore emphasizes the need for not just knowledge creation to promote acceptance of IRS but also the need for behavioral change communication and reinforcement during IRS campaigns.

### Conclusion

This study found that Indoor Residual Spraying in an urban community of an area that has holoendemic transmission of malaria covered only about half of the households. The factors associated with acceptance of IRS included younger age group below 35 years compared to those aged 36 years and older, those in formal employment compared to those in informal employment and those with moderate and good attitudes towards IRS. On the other hand, sex, marital status and educational levels if the heads of the households were not statistically associated with acceptance of IRS implementation in their households.

#### Recommendations

This study therefore recommends interventions that are aimed at changing the attitudes of people residing in high malaria transmission settings towards acceptance of IRS implementation especially people who are not employed and older people who may have entrenched misconceptions about the intervention.

## Data Availability

All data produced in the present study are available upon reasonable request to the authors

